# Association of subcutaneous or intravenous route of administration of casirivimab and imdevimab monoclonal antibodies with clinical outcomes in COVID-19

**DOI:** 10.1101/2021.11.30.21266756

**Authors:** Erin K. McCreary, J. Ryan Bariola, Richard J. Wadas, Judith A. Shovel, Mary Kay Wisniewski, Michelle Adam, Debbie Albin, Tami Minnier, Mark Schmidhofer, Russell Meyers, Oscar C. Marroquin, Kevin Collins, William Garrard, Lindsay R. Berry, Scott Berry, Amy M. Crawford, Anna McGlothlin, Kelsey Linstrum, Anna Nakayama, Stephanie K. Montgomery, Graham M. Snyder, Donald M. Yealy, Derek C. Angus, Paula L. Kip, Christopher W. Seymour, David T. Huang, Kevin E. Kip

**Author notes:** **Corresponding Author**: Erin K. McCreary, PharmD, BCPS, BCIDP, Clinical Assistant Professor of Medicine, University of Pittsburgh, Director of Stewardship Innovation, UPMC and Infectious Disease Connect, Falk Medical Building, Suite 5B, 3601 Fifth Avenue, Pittsburgh, PA 15213, C, E. **Funding:** This work did not receive external funding. The U.S. federal government provided the monoclonal antibody treatment reported in this manuscript.

## Abstract

**Importance:** Monoclonal antibody (mAb) treatment decreases hospitalization and death in outpatients with mild to moderate COVID-19; however, only intravenous administration has been evaluated in randomized clinical trials of treatment. Subcutaneous administration may expand outpatient treatment capacity and qualified staff available to administer treatment, but association with patient outcomes is understudied.

**Objective:** To evaluate whether or not, i.) subcutaneous casirivimab and imdevimab treatment is associated with reduced 28-day hospitalization/death than non-treatment among mAb-eligible patients, and ii.) subcutaneous casirivimab and imdevimab treatment is clinically and statistically similar to intravenous casirivimab and imdevimab treatment.

**Design, Setting, and Participants:** Prospective cohort study of outpatients in a learning health system in the United States with mild to moderate COVID-19 symptoms from July 14 to October 26, 2021 who were eligible for mAb treatment under emergency use authorization. A nontreated control group of eligible patients was also selected.

**Intervention:** Subcutaneous injection or intravenous administration of the combined single dose of casirivimab 600mg and imdevimab 600mg.

**Main Outcomes and Measures:** The primary outcome was the 28-day adjusted risk ratio or adjusted risk difference for hospitalization or death. Secondary outcomes included 28-day adjusted risk ratios/differences of hospitalization, death, composite endpoint of ED admission and hospitalization, and rates of adverse events.

**Results:** Among 1,956 matched adults with mild to moderate COVID-19, patients who received casirivimab and imdevimab subcutaneously had a 28-day rate of hospitalization/death of 3.4% (n=652) compared to 7.8% (n=1,304) in nontreated controls [risk ratio 0.44 (95% confidence interval: 0.28 to 0.68, p < .001)]. Among 2,185 patients treated with subcutaneous (n=969) or intravenous (n=1,216) casirivimab and imdevimab, the 28-day rate of hospitalization/death was 2.8% vs. 1.7%, respectively which resulted in an adjusted risk difference of 1.5% (95% confidence interval: -0.5% to 3.5%, p=.14). The 28-day adjusted risk differences (subcutaneous – intravenous) for death, ICU admission, and mechanical ventilation were 0.3% or less, although the 95% confidence intervals were wide.

**Conclusions and Relevance:** Subcutaneously administered casirivimab-imdevimab is associated with reduced risk-adjusted hospitalization or death amongst outpatients with mild to moderate COVID-19 compared to no treatment and indicates low adjusted risk difference compared to patients treated intravenously.

**Key Points:** *Question:* Among outpatients with mild to moderate COVID-19, is subcutaneously administered casirivimab and imdevimab associated with improved risk-adjusted 28-day clinical outcomes compared to non-treatment with monoclonal antibodies, and clinically similar association compared to intravenously administered casirivimab and imdevimab?

*Findings:* Among 1,956 propensity-matched adults, outpatients who received casirivimab and imdevimab subcutaneously had a 28-day rate of hospitalization or death of 3.4% (n=652) compared to 7.8% (n=1,304) in non-treated controls [risk ratio 0.44 (95% confidence interval: 0.28 to 0.68, p < .001)]. Among 2,185 outpatients who received subcutaneous (n=969) or intravenous (n=1,216) casirivimab and imdevimab, the 28-day rate of hospitalization/death was 2.8% vs. 1.7%, respectively, which resulted in an adjusted risk difference of 1.5% (95% confidence interval: -0.5% to 3.5%, p=.14). The 28-day adjusted risk differences comparing subcutaneous to intravenous route for death, ICU admission, and mechanical ventilation were 0.3% or less, although the 95% confidence intervals were wide.

*Meaning:* Subcutaneously administered casirivimab and imdevimab is associated with reduced hospitalization or death amongst outpatients with mild to moderate COVID-19 compared to no treatment, and has a small, adjusted risk difference compared to patients treated intravenously.

## INTRODUCTION

Discovery and broad-scale implementation of therapies that decrease progression to severe coronavirus disease 2019 (COVID-19) and improve mortality of patients infected with severe acute respiratory syndrome coronavirus 2 (SARS-CoV-2) are critical for global health. Casirivimab and imdevimab is a monoclonal antibody (mAb) that decreases hospitalizations and death in outpatients with mild to moderate COVID-19 when used as treatment or post-exposure prophylaxis.^1,2^ It is available under emergency use authorization (EUA) for these indications in the United States (US), United Kingdom (UK), and other global communities.^3,4^ Only intravenous administration was evaluated in randomized, clinical trials for treatment and accordingly, intravenous infusion is strongly recommended per the U.S. Food and Drug Administration (FDA) for this indication. However, the EUA states subcutaneous injection is an alternative route of administration when intravenous infusion is not feasible and would lead to delay in treatment, though efficacy of subcutaneous injection for treatment of SARS-CoV-2 is unknown.

A COVID-19 surge in September 2021, coupled with healthcare worker staffing shortages, resulted in a capacity crisis for outpatient monoclonal antibody infusions at our learning health system. Key stakeholders and clinical leaders determined that continuation of intravenous therapy would delay or prevent treatment for monoclonal antibody referrals, and conversion to subcutaneous injections would add treatment capacity, reduce appointment times, and expand staff available to administer treatment. The purpose of this study was to evaluate whether or not subcutaneous casirivimab and imdevimab is associated with reduced risk-adjusted 28-day clinical outcomes compared to non-treatment with monoclonal antibodies. We also sought to evaluate the similarity of clinical outcomes comparing subcutaneous with intravenous treatment to inform future operations within our learning health system.

## METHODS

This was a prospective cohort study of patients within the OPtimizing Treatment and Impact of Monoclonal antIbodieS Through Evaluation for COVID-19 embedded learning platform (OPTIMISE-C19, NCT04790786).^5^ This study was approved by the UPMC Quality Improvement Review Committee (Project ID 3629) and University of Pittsburgh Institutional Review Board (STUDY21100151). From platform launch on March 10, 2021 through September 9, 2021, all patients were assigned intravenous mAb treatment via a central management system. A small minority of patients received casirivimab and imdevimab subcutaneously if they presented directly to an urgent care facility within the system. From September 9 through October 26, 2021, most outpatient infusion centers provided only subcutaneous injections of casirivimab and imdevimab to accommodate surging patient referrals and staffing shortages. After October 26, centers converted back to intravenous administration when feasible within workforce capacity. Starting on September 28, 2021, patients aged 65 years or older with loss of two or more activities of daily living, pregnant patients, and/or patients with immunocompromised conditions were given priority for mAb treatment appointment scheduling.

### Outcomes

For this analysis, the two research questions were (1) whether or not subcutaneous casirivimab and imdevimab treatment is associated with better 28-day clinical outcomes than non-treatment among mAb-eligible patients; and (2) whether or not subcutaneous casirivimab and imdevimab treatment is clinically and statistically similar to intravenous casirivimab and imdevimab treatment. The primary outcome was the 28-day adjusted risk ratio of hospitalization/death for question 1, and the 28-day adjusted risk difference of hospitalization/death for question 2. Secondary outcomes included 28-day adjusted risk ratios/differences of hospitalization, death, composite endpoint of ED admission and hospitalization, and rates of adverse events.

### Selection of Patient Analysis Cohorts

For the first research question, nontreated control subjects were selected from non-hospitalized patients ≥ 12 years of age who had a positive SARS-CoV-2 polymerase chain reaction or antigen test within our health system from July 14 to October 26, 2021. These patients, whose symptom status was unknown, had an EUA-eligible risk factor for progression to severe disease and no admission to the emergency department or hospital on the date of their positive SARS-CoV-2 test result (i.e., presumed not to be at imminent risk of hospitalization). July 14^th^, 2021 was chosen as the start date for this analysis as this was the first confirmed date, per national tracking data, that 100% of patients infected with COVID-19 in our system had the Delta variant; Delta remained the only regional variant until the end of the study period.^6,7^ Corresponding treated subjects were patients ≥ 12 years of age treated subcutaneously with casirivimab and imdevimab in an outpatient infusion center or urgent care facility during the same period as nontreated control subjects. Patients receiving mAb in the emergency department were excluded since subcutaneous route of administration was not used in that setting. Both groups required a 28-day follow-up period. For nontreated control subjects, the 28-day outcome ascertainment period started on the day after the SARS-CoV-2 test positive result. For treated subjects, the 28-day outcome ascertainment period started on the day of mAb treatment.

For the second research question, patients treated subcutaneously or intravenously at an outpatient infusion center or urgent care facility on or after July 14, 2021 and with available follow-up period of 28 days were compared. For both groups, the 28-day outcome ascertainment period started on the day of treatment. Since not all clinical sites provided subcutaneous mAb treatment, separate study populations were compiled including all mAb treated patients, as well as the subset of mAb treated patients at clinical sites in which both routes of administration were used (i.e., to remove a potential site effect from the larger analysis). For this cohort to simply examine a scheduled change in practice, intravenous route patients were treated from July 15 to September 8, 2021 and subcutaneous route patients were treated from September 9 to September 29, 2021 (i.e., non-overlapping treatment periods).

### Data Sources

We used health-related data captured in the electronic health record and ancillary clinical systems, all of which are aggregated and harmonized in a Clinical Data Warehouse (CDW).^8,9^ For infusion sites with complete EMR data in the CDW, we accessed sociodemographic data, medical history, and billing charges for all outpatient and in-hospital encounters with diagnoses and procedures coded based on the International Classification of Diseases, Ninth and Tenth revisions (ICD-9 and ICD-10, respectively).^10,11^ We assessed 28-day mortality by the hospital discharge disposition of “Ceased to Breathe” sourced from the inpatient Medical Record System, as well as deaths after discharge identified with the Death Master File from the Social Security Administration, 2021 National Technical Information Service as an external data source.^12,13^

### Statistical Methods

Sociodemographic and clinical characteristics were compared between subjects treated subcutaneously and nontreated control subjects by use of student *t*-tests or Wilcoxon tests for continuous variables (depending on distributional properties) and chi-square tests for categorical variables. To control for imbalances in patient profiles between the two groups, we selected nontreated control subjects matched to treated subjects by propensity score methodology.^14,15^ Specifically, propensity scores were derived from a logistic regression model fit from a multitude of variables with subcutaneous mAb treatment as the response variable and forward stepwise selection of measured pre-treatment explanatory variables at p <0.15. We included the variables age, gender, and race into the model prior to stepwise selection. We used 1:2 propensity score matching with a maximum propensity score probability difference of 0.01 to construct the matched treated and nontreated groups. We performed non-matched parallel analyses in which outcomes of treated subjects were compared to nontreated subjects using (adjusting for) the propensity score as a covariate, and with inverse probability weighting. Both the matched and non-matched adjusted analyses were conducted using generalized linear models with mAb receipt as the predictor of interest, specifying the binomial distribution and log link. We did not impute missing values for variables used in deriving the propensity scores.

Sociodemographic and clinical characteristics were also compared between subjects treated subcutaneously and intravenously by use of student *t*-tests or Wilcoxon tests for continuous variables and chi-square tests for categorical variables. Given few between-group patient differences, we used stepwise logistic regression to examine potential confounders, with age, gender, and vaccination status included as covariates in all models. The primary parameter of interest was the adjusted risk difference (subcutaneous – intravenous) in the 28-day rate of hospitalization/death with a boundary of 3% used to define similar clinical outcome. A 3% boundary was decided as a consensus threshold amongst authors as clinically meaningful for the health system population and for capacity management.

All analyses were performed using the SAS System (Cary, NC), version 9.4. Methods and results are reported in accordance with The REporting of studies Conducted using Observational Routinely-Collected health Data (RECORD) statement (**eTable 1**).^16^

## RESULTS

### Study Populations

For the first analysis, there were 969 patients treated subcutaneously with casirivimab and imdevimab and 4,353 non-treated, EUA-eligible controls in the unmatched cohort. The propensity score-matched analysis, which required complete covariate data and matching (as defined in methods), compared 652 patients treated subcutaneously with casirivimab and imdevimab to 1,304 non-treated, EUA-eligible controls (**Figure 1**). For the second analysis, 969 patients treated subcutaneously with casirivimab and imdevimab were compared to 1,216 patients treated with the same mAb intravenously, of whom, 721 versus 441 patients were treated at clinical sites in which both routes of administration were used during the study period (**Figure 1**).

**Figure 1.**
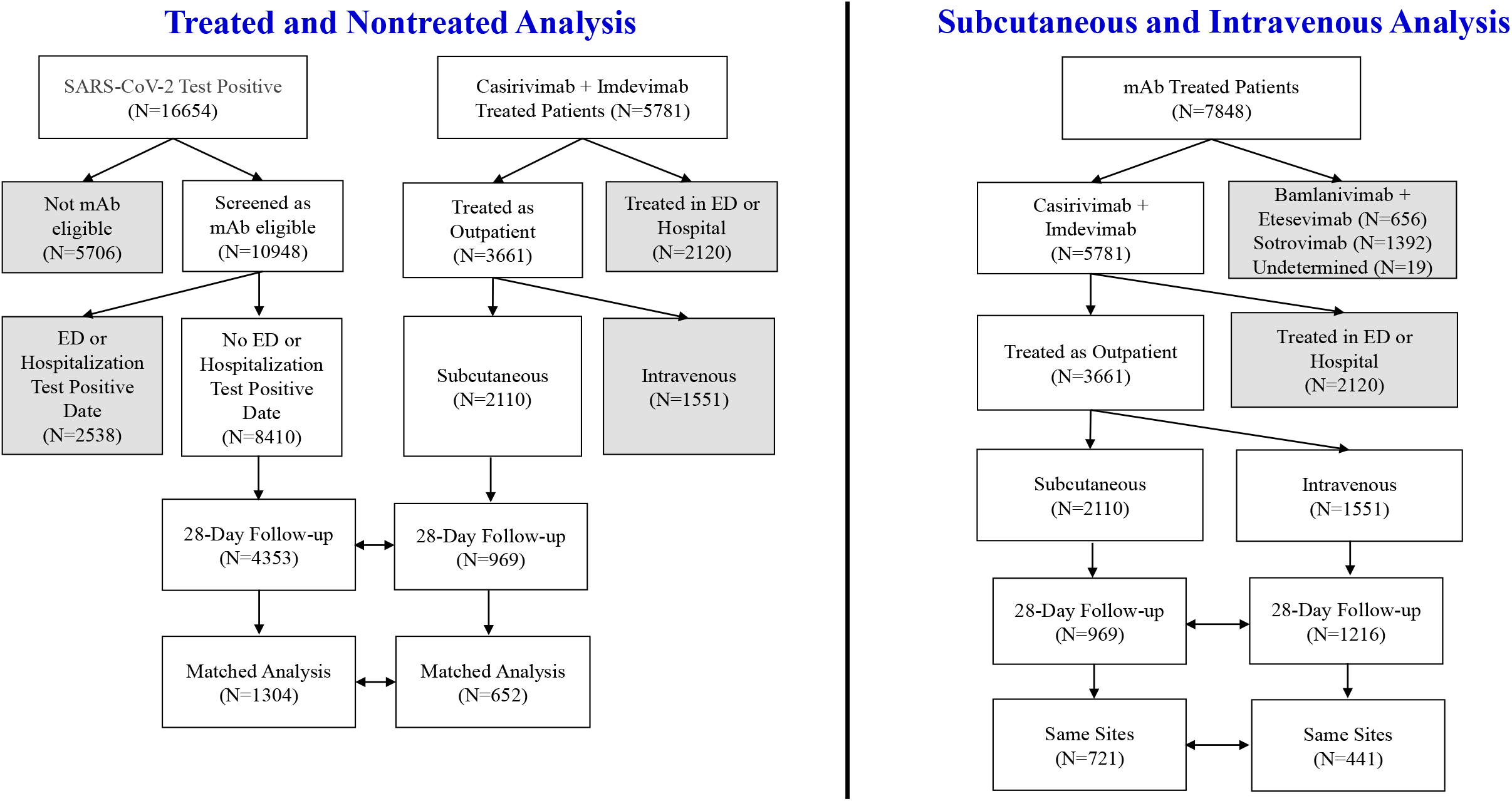
CONSORT diagram of the selection criteria and patient populations used in the superiority analysis of subcutaneous treatment with C + I versus nontreated control subjects (left side of diagram), and the non-equivalence analysis of subcutaneous and intravenous treatment (right side of the diagram). Shaded boxes depict subjects excluded from analysis.

### Matched Analysis of Treated and Nontreated Patients

Prior to matching on propensity score (based on covariate data), subcutaneously treated patients (who were selected using a priority system favoring older age and being immunocompromised starting on September 28, 2021) were significantly older and less likely of Black race than nontreated patients (**Table 1**). In addition, treated patients had a significantly higher prevalence of rheumatoid arthritis, obstructive sleep apnea, hypertension, and smoking history than nontreated patients. This overall higher risk profile of treated patients was also reflected in a higher prevalence of statin and beta blocker use than nontreated patients. Importantly, after propensity score matching, treated and nontreated patients were generally similar on variables included in the propensity score model (**Table 1, upper portion**), the distribution of propensity scores (**eFigure 1**), and variables not included in the model (**Table 1, lower portion**).

**Table 1.** Comparison of Characteristics in Subcutaneous mAb Treated Group and Nontreated Control Group.

| Characteristic | Unmatched |  |  | Matched |  |  |
| --- | --- | --- | --- | --- | --- | --- |
|  | Treated<br>(N=969) | Nontreated<br>(N=4353) | p-value | Treated<br>(N=652) | Nontreated<br>(N=1304) | p-value |
| Age, mean (SD) | 53.8 (16.7) | 49.7 (21.5) | <.001 | 53.7 (16.9) | 53.0 (19.3) | .44 |
| Female gender, No. (%) | 547 (56.4) | 2542 (58.4) | .27 | 388 (59.5) | 785 (60.2) | .77 |
| Black race, No. (%) | 49 (5.2) | 407 (9.5) | <.001 | 35 (5.4) | 50 (3.8) | .12 |
| Corticosteroids as a home medication, No. (%) | 240 (35.9) | 1190 (30.9) | .01 | 231 (35.4) | 448 (34.4) | .64 |
| History of rheumatoid arthritis, No. (%) | 24 (3.6) | 62 (1.6) | <.001 | 20 (3.1) | 30 (2.3) | .31 |
| History of asthma, No. (%) | 220 (32.9) | 1395 (36.2) | .10 | 213 (32.7) | 387 (29.7) | .18 |
| History of obstructive sleep apnea, No. (%) | 128 (19.2) | 507 (13.2) | <.001 | 123 (18.9) | 212 (16.3) | .15 |
| History of hypertension, No. (%) | 314 (47.0) | 1422 (36.9) | <.001 | 303 (46.5) | 591 (45.3) | .63 |
| History of COPD, No. (%) | 115 (17.2) | 566 (14.7) | .09 | 108 (16.6) | 202 (15.5) | .54 |
| History of diabetes, No. (%) | 112 (16.8) | 559 (14.5) | .13 | 107 (16.4) | 203 (15.6) | .63 |
| History of solid organ or cell transplant, No. (%) | 10 (1.5) | 6 (0.2) | <.001 | 1 (0.2) | 2 (0.2) | 1.0 |
| **Variables Not Included in Propensity Score** | -- | -- | -- | -- | -- | -- |
| History of smoking, No. (%) | 227 (34.0) | 777 (28.7) | .008 | 221 (33.9) | 305 (33.5) | .88 |
| Body mass index, mean (SD) | 31.8 (7.5) | 31.6 (7.7) | .47 | 32.0 (7.6) | 32.1 (7.7) | .92 |
| History of dyspnea, No. (%) | 40 (6.0) | 200 (5.2) | .40 | 40 (6.1) | 75 (5.7) | .52 |
| History of pulmonary hypertension, No. (%) | 13 (1.9) | 56 (1.4) | .34 | 11 (1.7) | 21 (1.6) | .90 |
| History of atrial fibrillation, No. (%) | 33 (4.9) | 167 (4.3) | .48 | 32 (4.9) | 62 (4.7) | .88 |
| History of valvular heart disease, No. (%) | 31 (4.6) | 178 (4.6) | .98 | 30 (4.6) | 71 (5.4) | .43 |
| History of coronary artery disease, No. (%) | 73 (10.9) | 320 (8.3) | .03 | 71 (10.9) | 142 (10.9) | 1.0 |
| History of stroke, No. (%) | 35 (5.2) | 185 (4.8) | .63 | 33 (5.1) | 84 (6.4) | .22 |
| History of congestive heart failure, No. (%) | 36 (5.4) | 221 (5.7) | .72 | 32 (4.9) | 85 (6.5) | .16 |
| History of chronic kidney disease, No. (%) | 34 (5.1) | 185 (4.8) | .75 | 27 (4.1) | 80 (6.1) | .07 |
| History of fatty liver disease, No. (%) | 28 (4.2) | 106 (2.8) | .04 | 28 (4.3) | 42 (3.2) | .23 |
| History of cancer, No. (%) | 70 (10.5) | 310 (8.0) | .04 | 69 (10.6) | 133 (10.2) | .79 |
| History of chemotherapy, No. (%) | 31 (4.6) | 113 (2.9) | .02 | 28 (4.3) | 37 (2.8) | .09 |
| History of allergic rhinitis, No. (%) | 78 (11.7) | 517 (13.4) | .22 | 77 (11.8) | 172 (13.2) | .39 |
| History of viral hepatitis, No. | 12 (1.8) | 44 (1.1) | .16 | 11 (1.7) | 19 (1.5) | .70 |
| (%) |  |  |  |  |  |  |
| ACE Inhibitors, No. (%) | 118 (17.7) | 525 (13.6) | .006 | 118 (18.1) | 201 (15.4) | .13 |
| Angiotensin II receptor blocker, No. (%) | 69 (10.3) | 316 (8.2) | .07 | 67 (10.3) | 152 (11.7) | .36 |
| Alpha blocker, No. (%) | 9 (1.3) | 44 (1.1) | .65 | 9 (1.4) | 16 (1.2) | .78 |
| Beta blockers, No. (%) | 166 (24.8) | 708 (18.4) | <.001 | 158 (24.2) | 272 (20.9) | .09 |
| Statins, No. (%) | 223 (33.4) | 1018 (26.4) | <.001 | 216 (33.1) | 425 (32.6) | .81 |
| Antidepressants, No. (%) | 200 (29.9) | 1115 (28.9) | .60 | 194 (29.7) | 414 (31.7) | .37 |
| Charlson Comorbidity Index Score, mean (SD) | 0.8 (1.4) | 0.7 (1.3) | .03 | 0.8 (1.4) | 0.8 (1.4) | .96 |

Abbreviations: SD, standard deviation; COPD, chronic obstructive pulmonary disease; ACE, angiotensin - converting enzyme.

The matched 28-day rate of hospitalization/death was 3.4% in treated patients compared to 7.8% in nontreated controls (**Table 2**). The corresponding risk ratio for 28-day hospitalization/death was 0.44 (95% confidence interval: 0.28 to 0.68, p < .001). The lower risk of hospitalization/death in treated patients was most evident in the first 15 days of follow-up (**eFigure 2**). The 28-day death rate was 0.2% in the treated group vs. 2.2% in the nontreated group (p=.009 from the log-binomial regression model).

**Table 2.** Propensity Matched 28-Day Event Free Rates and Risk Ratios of Study Outcomes.

| **28-Day Event Free Outcomes** | **Number of Events** |  | **28-Day Event Rate (%)** |  | **Risk Ratio Estimates** |  |  |
| --- | --- | --- | --- | --- | --- | --- | --- |
|  | **Treated (n=652)** | **Nontreated (n=1304)** | **Treated** | **Nontreated** | **Risk Ratio** | **(95% CI)** | **p-value** |
| Hospitalization/death | 22 | 101 | 3.4 | 7.8 | 0.44 | (0.28 – 0.68) | <.001 |
| Hospitalization | 22 | 85 | 3.4 | 6.5 | 0.52 | (0.33 – 0.82) | .005 |
| Death | 1 | 29 | 0.2 | 2.2 | 0.07 | (0.01 – 0.51) | .009 |
| ED admission or hospitalization | 40 | 133 | 6.1 | 10.2 | 0.60 | (0.43 – 0.85) | .003 |

Abbreviations: CI, confidence interval; ED, emergency department.

### Unmatched Analysis of Treated and Nontreated Patients

In unmatched patients with a propensity score (i.e., covariate data), the crude 28-day rate of hospitalization/death was 3.5% in treated patients compared to 6.6% in nontreated controls (**eTable 2**). The corresponding risk ratio for hospitalization or death adjusted for the propensity score was 0.44 (95% confidence interval: 0.29 to 0.66, p < .001). Results were relatively consistent with the use of inverse probability weighting. As in the matched analysis, deaths were infrequent with the 28-day death rate being lower in the treated group (0.2%) compared to the nontreated group (2.1%) (adjusted risk ratio = 0.06, 95% confidence interval: 0.01 to 0.41, p=.004).

### Evaluation of Subcutaneous and Intravenous Treatment

Patients treated subcutaneously or intravenously had a mean age of 54 years and a mean Charlson Comorbidity Index score of 0.8; the two groups were also generally similar on demographic and presenting clinical characteristics (**Table 3**). This overall similarity in patient profiles was evident among all treated patients as well as the subset of patients treated at clinical sites in which both routes of mAb administration were used. A notable exception was a higher rate of full COVID-19 vaccination in patients treated subcutaneously (55.5%) compared to those treated intravenously (44.1%) in patients from all sites; however, this rate was similar in patients treated within same sites. The median time (IQR) from symptom onset to infusion was 6 (5, 8) in both groups.

**Table 3.** Comparison of Characteristics of Subcutaneous and Intravenous Monoclonal Antibody Treated Patients.

| Characteristic | <u>All Infused Patients*</u> |  |  | <u>Same Site Infused Patients**</u> |  |  |
| --- | --- | --- | --- | --- | --- | --- |
|  | Subcutaneous | Intravenous |  | Subcutaneous | Intravenous |  |
|  | (N=969) | (N=1216) | p-value | (N=721) | (N=441) | p-value |
| Age, mean (SD) | 53.8 (16.7) | 54.3 (16.6) | .45 | 54.5 (16.5) | 53.9 (17.4) | .57 |
| Female gender, No. (%) | 547 (56.4) | 672 (54.4) | .35 | 401 (55.6) | 227 (51.5) | .17 |
| Race, No. (%) | -- | -- | .08 | -- | -- | .55 |
| White | 852 (89.8) | 1071 (89.4) | -- | 640 (89.1) | 392 (89.3) | -- |
| Black | 49 (5.2) | 83 (6.9) | -- | 32 (4.5) | 24 (5.5) | -- |
| Other | 48 (5.1) | 44 (3.7) | -- | 46 (6.4) | 23 (5.25) | -- |
| History of smoking, No. (%) | 227 (34.0) | 135 (31.2) | .35 | 171 (33.9) | 113 (31.0) | .46 |
| Body mass index, mean (SD) | 31.8 (7.5) | 32.8 (8.4) | .03 | 32.0 (7.8) | 32.3 (8.4) | .62 |
| History of diabetes, No. (%) | 112 (16.8) | 161 (17.1) | .86 | 93 (18.4) | 58 (16.2) | .39 |
| History of obstructive sleep apnea, No. (%) | 128 (19.2) | 174 (18.5) | .73 | 106 (21.0) | 76 (21.2) | .95 |
| History of dyspnea, No. (%) | 40 (6.0) | 69 (7.3) | .29 | 33 (6.5) | 17 (4.7) | .26 |
| History of asthma, No. (%) | 220 (32.9) | 283 (30.0) | .22 | 164 (32.5) | 114 (31.7) | .82 |
| History of pulmonary hypertension, No. (%) | 13 (1.9) | 9 (1.0) | .09 | 11 (2.2) | 1 (0.3) | .02 |
| History of COPD, No. (%) | 115 (17.2) | 151 (16.0) | .53 | 84 (16.6) | 54 (15.0) | .53 |
| History of hypertension, No. (%) | 314 (47.0) | 408 (43.3) | .14 | 253 (50.1) | 158 (44.0) | .08 |
| History of atrial fibrillation, No. (%) | 33 (4.9) | 55 (5.8) | .43 | 27 (5.4) | 20 (5.6) | .87 |
| History of valvular heart disease, No. (%) | 31 (4.6) | 68 (7.2) | .03 | 26 (5.2) | 29 (8.1) | .08 |
| History of coronary artery disease, No. (%) | 73 (10.9) | 105 (11.1) | .89 | 61 (12.1) | 45 (12.5) | .84 |
| History of stroke, No. (%) | 35 (5.2) | 37 (3.9) | .21 | 28 (5.5) | 16 (4.5) | .47 |
| History of congestive heart failure, No. (%) | 36 (5.4) | 50 (5.3) | .94 | 26 (5.1) | 13 (3.6) | .29 |
| History of chronic kidney disease, No. (%) | 34 (5.1) | 63 (6.7) | .18 | 24 (4.7) | 24 (6.7) | .22 |
| History of fatty liver disease, No. (%) | 28 (4.2) | 23 (2.4) | .05 | 23 (4.5) | 11 (3.1) | .27 |
| History of cancer, No. (%) | 70 (10.5) | 127 (13.5) | .07 | 53 (10.5) | 57 (15.9) | .02 |
| History of chemotherapy, No. (%) | 31 (4.6) | 35 (3.7) | .36 | 26 (5.1) | 26 (7.2) | .20 |
| History of allergic rhinitis, No. (%) | 78 (11.7) | 140 (14.9) | .07 | 56 (11.1) | 47 (13.1) | .37 |
| History of rheumatoid arthritis, (No.), % | 24 (3.6) | 26 (2.8) | .34 | 17 (3.4) | 15 (4.2) | .53 |
| History of viral hepatitis, No. (%) | 12 (1.8) | 10 (1.1) | .21 | 5 (1.0) | 2 (0.6) | .48 |
| History of solid organ or cell transplant, No. (%) | 10 (1.5) | 10 (1.1) | .44 | 4 (0.8) | 7 (1.9) | .22 |
| ACE Inhibitors, No. (%) | 118 (17.7) | 138 (14.7) | .10 | 105 (20.8) | 52 (14.5) | .02 |
| Angiotensin II receptor blocker, No. (%) | 69 (10.3) | 102 (10.8) | .75 | 53 (10.5) | 45 (12.5) | .35 |
| Alpha blocker, No. (%) | 9 (1.3) | 7 (0.7) | .23 | 6 (1.2) | 1 (0.3) | .14 |
| Beta blockers, No. (%) | 166 (24.8) | 198 (21.0) | .07 | 126 (24.9) | 89 (24.8) | .96 |
| Statins, No. (%) | 223 (33.4) | 333 (35.4) | .41 | 181 (35.8) | 134 (37.3) | .66 |
| Antidepressants, No. (%) | 200 (29.9) | 314 (33.3) | .15 | 153 (30.3) | 132 (36.8) | .05 |
| Corticosteroids as a home medication, No. (%) | 240 (35.9) | 340 (36.1) | .95 | 181 (35.8) | 114 (31.8) | .21 |
| Charlson Comorbidity Index Score, mean (SD) | 0.8 (1.4) | 0.8 (1.3) | .95 | 0.8 (1.4) | 0.8 (1.2) | .75 |
| Days from symptoms to infusion, mean (SD) | 6.1 (1.9) | 6.1 (2.0) | .95 | 6.1 (1.9) | 6.1 (1.9) | .83 |
| Days from symptoms to infusion, No. (%) | -- | -- | .11 | -- | -- | .73 |
| 1 to 4 | 170 (21.2) | 257 (23.4) | -- | 144 (21.0) | 85 (20.9) | -- |
| 5 to 6 | 293 (36.5) | 352 (32.0) | -- | 248 (36.3) | 139 (34.1) | -- |
| 7 or more | 339 (42.3) | 491 (44.6) | -- | 292 (42.7) | 183 (45.0) | -- |
| Vaccination status, No. (%) | -- | -- | <.001 | -- | -- | .75 |
| Unvaccinated | 267 (33.2) | 455 (41.4) | -- | 232 (33.8) | 141 (34.6) | -- |
| Partially vaccinated | 22 (2.7) | 26 (2.4) | -- | 18 (2.6) | 14 (3.4) | -- |
| Fully vaccinated | 447 (55.5) | 485 (44.1) | -- | 372 (54.2) | 210 (51.6) | -- |
| Unknown/not determined | 69 (8.6) | 134 (12.2) | -- | 64 (9.3) | 42 (10.3) | -- |

Abbreviations: SD, standard deviation; COPD, chronic obstructive pulmonary disease; ACE, angiotensin - converting enzyme.
\*Patients treated at all health system facilities; subcutaneous patients were treated from July 20 – September 29, 2021; intravenous patients were treated from July 15- September 29, 2021.
\*\*Patients treated at the same health system facilities; subcutaneous patients were treated from September 9-29, 2021; intravenous patients were treated from July 15- September 8, 2021.

For all infused patients, the adjusted risk difference of hospitalization/death comparing subcutaneous and intravenous treated patients was 1.5% (95% confidence interval: -0.5% to 3.5%, p=.14) which was within the clinically predefined similarity boundary of 3%, yet the upper limit of the 95% confidence interval marginally exceeded this boundary (**Table 4**). The corresponding adjusted risk ratio was 1.71 (95% confidence interval: 0.97 to 3.00, p=.06). Adjusted risk differences of death and ED admission/hospitalization were small (similar) by route of administration. In terms of initial safety, rates of severe adverse reactions comparing subcutaneous to intravenous patients were 0.0% and 0.2%, respectively.

**Table 4.**
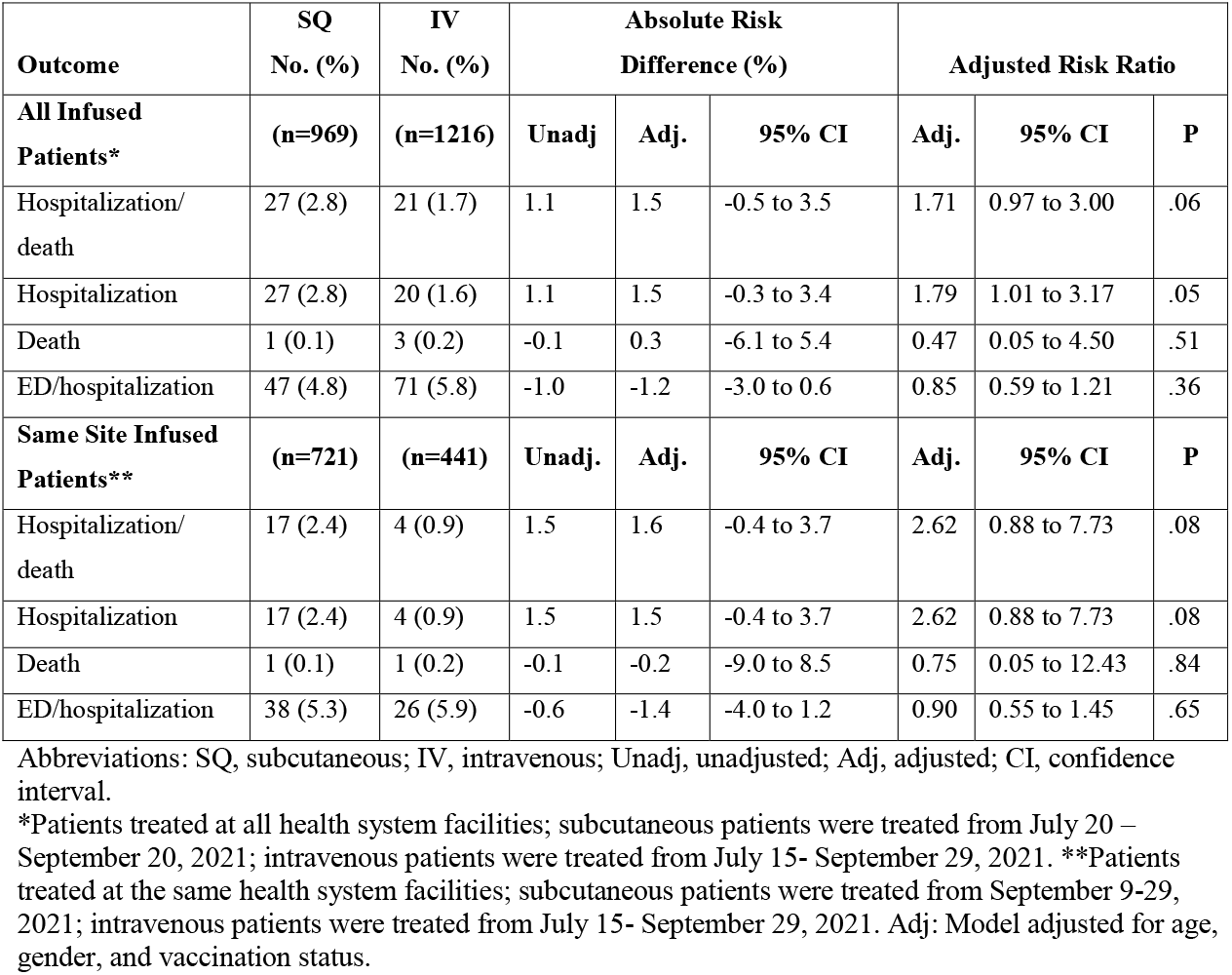
28-Day Risk Differences and Risk Ratios of Hospitalization or Death by Route of Monoclonal Antibody Administration.

| Outcome | SQ | IV | Absolute Risk |  |  | Adjusted Risk Ratio |  |  |
| --- | --- | --- | --- | --- | --- | --- | --- | --- |
|  | No. (%) | No. (%) | Difference (%) |  |  |  |  |  |
| **All Infused Patients\*** | **(n=969)** | **(n=1216)** | **Unadj** | **Adj.** | **95% CI** | **Adj.** | **95% CI** | **P** |
| Hospitalization/death | 27 (2.8) | 21 (1.7) | 1.1 | 1.5 | -0.5 to 3.5 | 1.71 | 0.97 to 3.00 | .06 |
| Hospitalization | 27 (2.8) | 20 (1.6) | 1.1 | 1.5 | -0.3 to 3.4 | 1.79 | 1.01 to 3.17 | .05 |
| Death | 1 (0.1) | 3 (0.2) | -0.1 | 0.3 | -6.1 to 5.4 | 0.47 | 0.05 to 4.50 | .51 |
| ED/hospitalization | 47 (4.8) | 71 (5.8) | -1.0 | -1.2 | -3.0 to 0.6 | 0.85 | 0.59 to 1.21 | .36 |
| **Same Site Infused Patients\*\*** | **(n=721)** | **(n=441)** | **Unadj.** | **Adj.** | **95% CI** | **Adj.** | **95% CI** | **P** |
| Hospitalization/death | 17 (2.4) | 4 (0.9) | 1.5 | 1.6 | -0.4 to 3.7 | 2.62 | 0.88 to 7.73 | .08 |
| Hospitalization | 17 (2.4) | 4 (0.9) | 1.5 | 1.5 | -0.4 to 3.7 | 2.62 | 0.88 to 7.73 | .08 |
| Death | 1 (0.1) | 1 (0.2) | -0.1 | -0.2 | -9.0 to 8.5 | 0.75 | 0.05 to 12.43 | .84 |
| ED/hospitalization | 38 (5.3) | 26 (5.9) | -0.6 | -1.4 | -4.0 to 1.2 | 0.90 | 0.55 to 1.45 | .65 |

Abbreviations: SQ, subcutaneous; IV, intravenous; Unadj, unadjusted; Adj, adjusted; CI, confidence interval.
\*Patients treated at all health system facilities; subcutaneous patients were treated from July 20 – September 20, 2021; intravenous patients were treated from July 15- September 29, 2021. \*\*Patients treated at the same health system facilities; subcutaneous patients were treated from September 9-29, 2021; intravenous patients were treated from July 15- September 29, 2021. Adj: Model adjusted for age, gender, and vaccination status.

Among patients treated at clinical sites in which both routes of administration were used, the 28-day risk difference (subcutaneous – intravenous) of hospitalization/death was 1.6% (95% confidence interval: -0.4% to 3.7%, p=.12) (**Table 4**). The corresponding adjusted risk ratio was 2.62 (95% confidence interval: 0.88 to 7.73, p=.08). Adjusted risk differences and risk ratios for death and ED admission/hospitalization were non-significantly lower in the direction favoring subcutaneous treated patients.

In supplemental analysis to investigate the clinically small (higher) 28-day risk difference of hospitalization/death in patients treated subcutaneously, the 28-day risk differences of ICU admission and mechanical ventilation (i.e., indicators of severity of hospitalization) were very low and similar in patients treated subcutaneously or intravenously (**eTable 3**). In addition, length of stay was similar by route of administration among hospitalized patients.

## Discussion

Among a matched analysis of 1,956 patients, subcutaneously administered casirivimab and imdevimab was associated with an estimated 56% lower risk of 28-day hospitalization/death compared to no mAb treatment amongst EUA-eligible outpatients. Among 2,185 patients with mild to moderate COVID-19 treated at an outpatient infusion center, the adjusted risk difference of 28-day hospitalization/death comparing subcutaneous and intravenous mAb treatment was 1.5%, below our pre-defined similarity boundary of 3%. However, the upper limit of the 95% confidence interval (3.5%) exceeded the clinical boundary indicating a possible increased risk of hospitalization with subcutaneous mAb administration. On the other hand, there was little to no evidence that subcutaneous administration was associated with a higher risk of death or severe hospitalization (i.e., ICU admission or mechanical ventilation). Collectively, these data suggest that subcutaneous administration of mAb may be a reasonable alternative to intravenous administration for prevention of death and severe hospitalization.

To our knowledge, this report is the largest analysis of outpatients with mild to moderate COVID-19 treated with subcutaneously administered mAb compared to nontreated and intravenously administered mAb. These non-causal data indicate a consistent, significant benefit of mAb therapy in decreasing hospitalizations and deaths for patients with mild to moderate COVID-19, regardless of route of administration in a 100% Delta variant landscape. The adjusted risk difference between subcutaneous and intravenous administration for rate of 28-day hospitalization or death was small and not statistically significant, and there was no difference in risk of severity of illness once hospitalized. This evidence is promising as administering intravenous monoclonal antibodies is logistically challenging, and health systems across the globe continue to face critical staffing shortages amidst high SARS-CoV-2 positive patient volumes. Subcutaneous administration of monoclonal antibodies allows for reduced appointment times (due to elimination of need to place a venous access line and need to infuse the medication over a certain number of minutes), which increases treatment capacity. Indeed, our health system was able to increase the number of patient appointments for mAb treatment from 400 to 1,000 patients per week with the same number of staff by changing the route of administration from intravenous to subcutaneous. Additionally, under the Public Readiness and Emergency Preparedness (PREP) Act in the US, pharmacists are allowed to administer subcutaneous injections, expanding the available staffing pool to much greater capacity.^17^ These important gains in practical resources for stressed health systems must be weighed against the absolute risk difference in hospitalizations with subcutaneous administration and intravenous administration, particularly when assessed in relation to lower risk of hospitalization and death for subcutaneous administration in patients compared to nontreated patients.

Access to safe and effective outpatient treatments for COVID-19 is of critical importance to the global community, and subcutaneous mAb administration has useful implications for scaling resources. By avoiding limitations associated with intravenous administration, subcutaneous mAb treatment and post-exposure prophylaxis outpatient treatment location sites can potentially reach disadvantaged neighborhoods and low middle-income countries more readily.

Our study has limitations. First, nontreated controls were matched by EUA-eligible risk factors only and we were unable to determine symptom severity (whether symptomatic or asymptomatic) in these patients. Thus, many nontreated patients may have been asymptomatic and thereby at low risk of hospitalization, which would tend to bias results against mAb treatment. Second, although adjusted for statistically, more patients in the subcutaneous group were fully vaccinated compared to the intravenous group at all sites, which may also lower risk of hospitalization and death. However, “fully vaccinated” on the referral form meant receipt of two doses of an mRNA vaccine or one dose of an adenovirus vaccine—further details on time from last dose to mAb referral, type of vaccine, or whether or not a third primary series dose had been administered to immunocompromised patients was unknown and therefore fully vaccinated cannot be interpreted as fully protected. This difference was also mitigated when the analysis was restricted to patients treated at same sites. Finally, the mean time from symptom onset to mAb treatment in our study was 6 days. While these therapies work best earliest in disease course, administering treatment faster in real-world settings is logistically challenging and the observed time to treatment in this study represents best practices for mAb treatment across an extensive geographical region. Time to treatment windows will be important to consider as novel, oral antiviral medications become available with reduced treatment windows compared to mAb treatment.^18,19^

## Conclusions

Subcutaneously administered casirivimab and imdevimab was associated with an estimated 56% lower risk of hospitalization or death as compared to no monoclonal antibody treatment in at-risk outpatients with mild to moderate COVID-19. The adjusted risk difference of hospitalization or death comparing subcutaneous and intravenous treated patients was 1.5% (95% confidence interval: -0.5% to 3.5%). Moreover, there was no difference in 28-day risk of death, ICU admission, or mechanical ventilation between subcutaneously or intravenously treated patients. Collectively, these results provide preliminary evidence for potential expanded use of subcutaneous mAb treatment, particularly in areas facing treatment capacity and/or staffing shortages.

## Data Availability

All data produced in the present study are available upon reasonable request to the authors.

---

**eTable 1.**
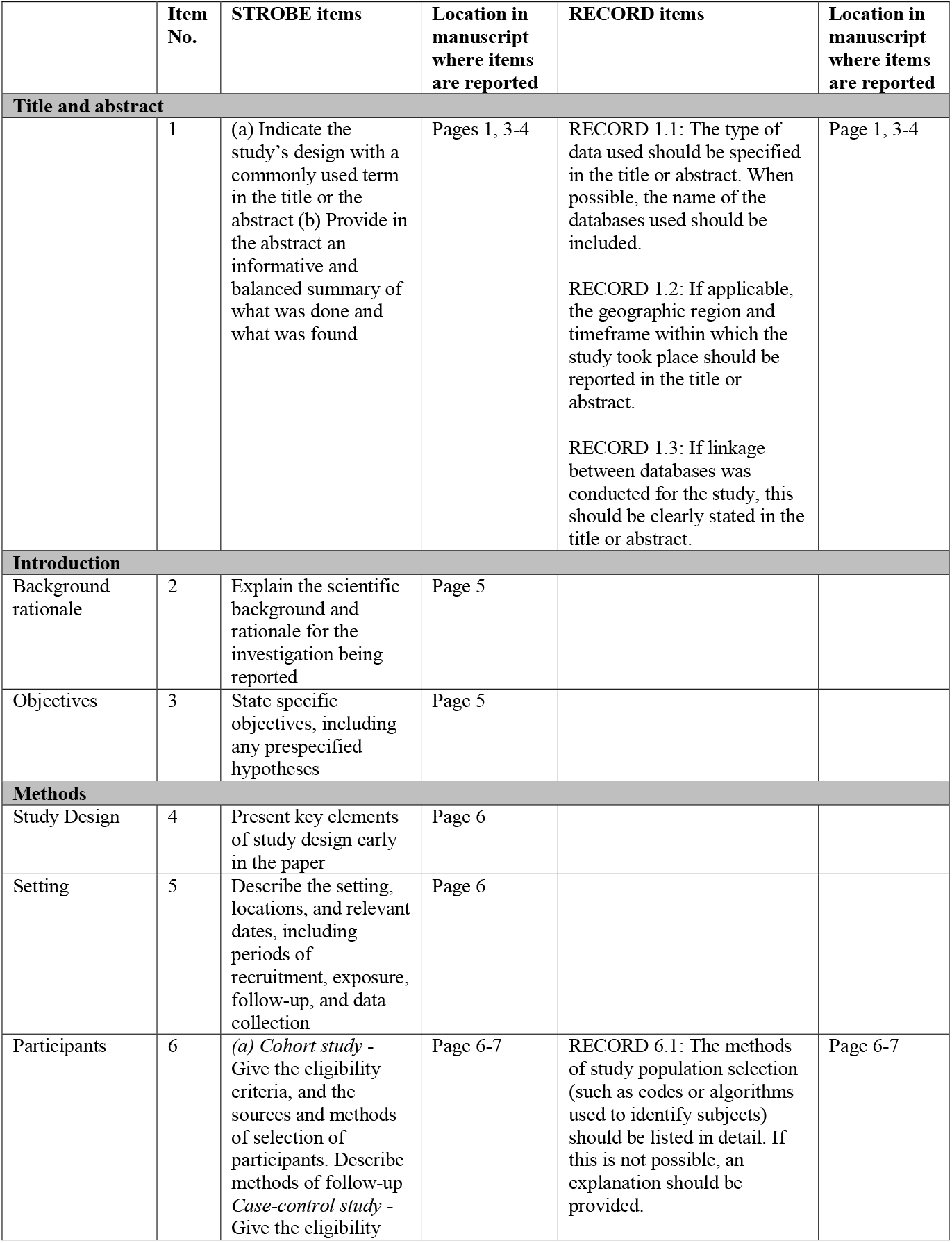
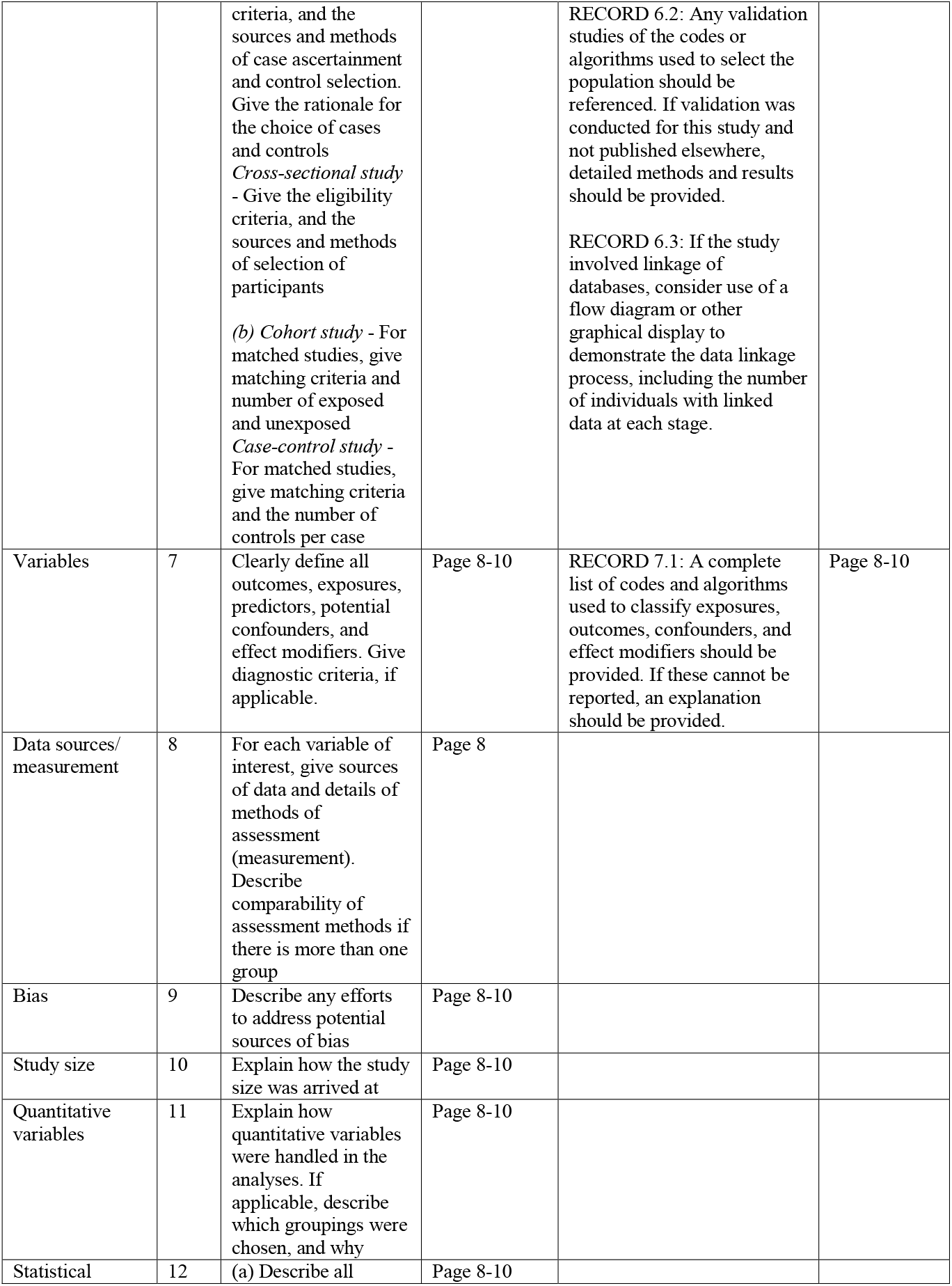
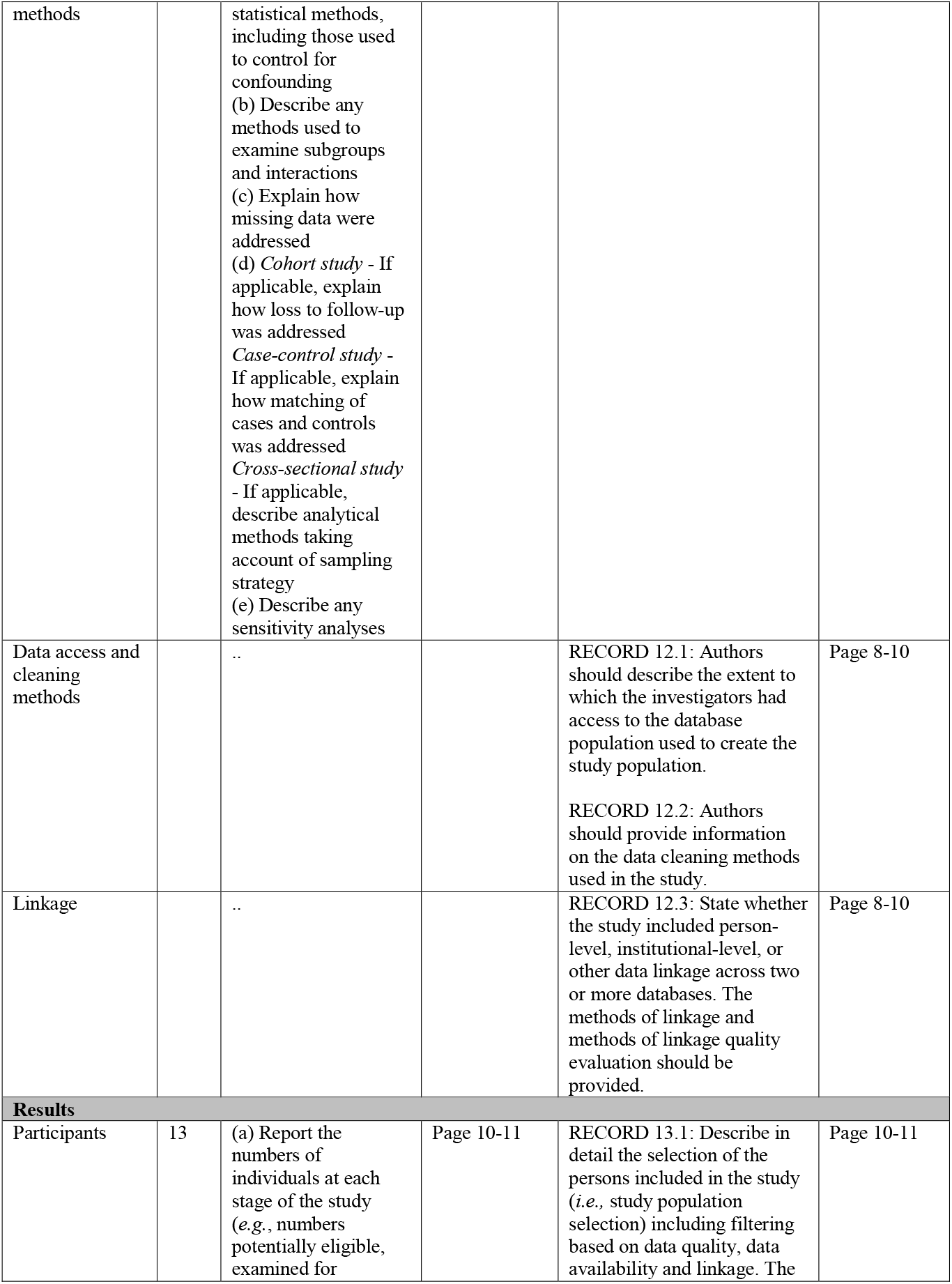
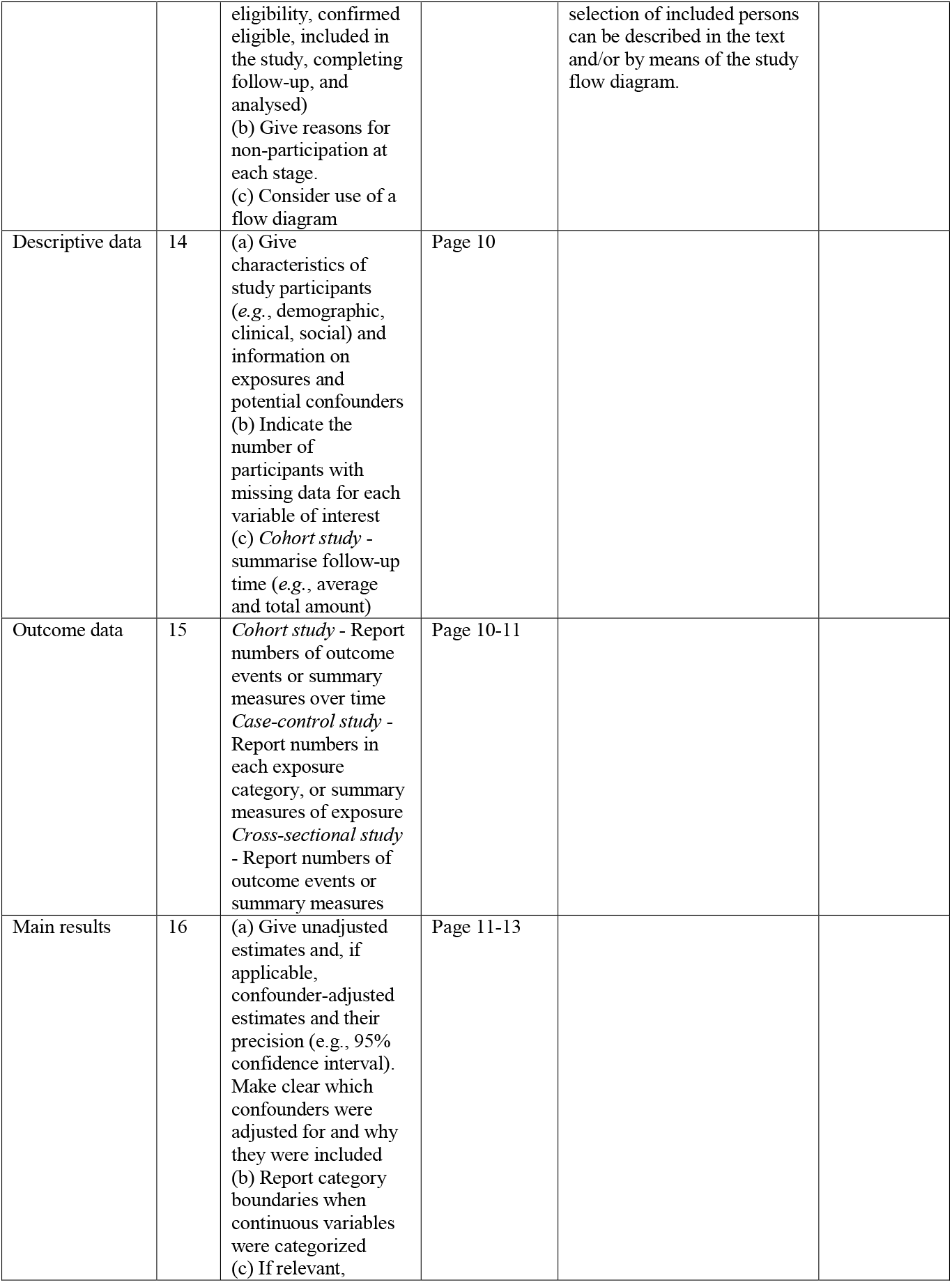
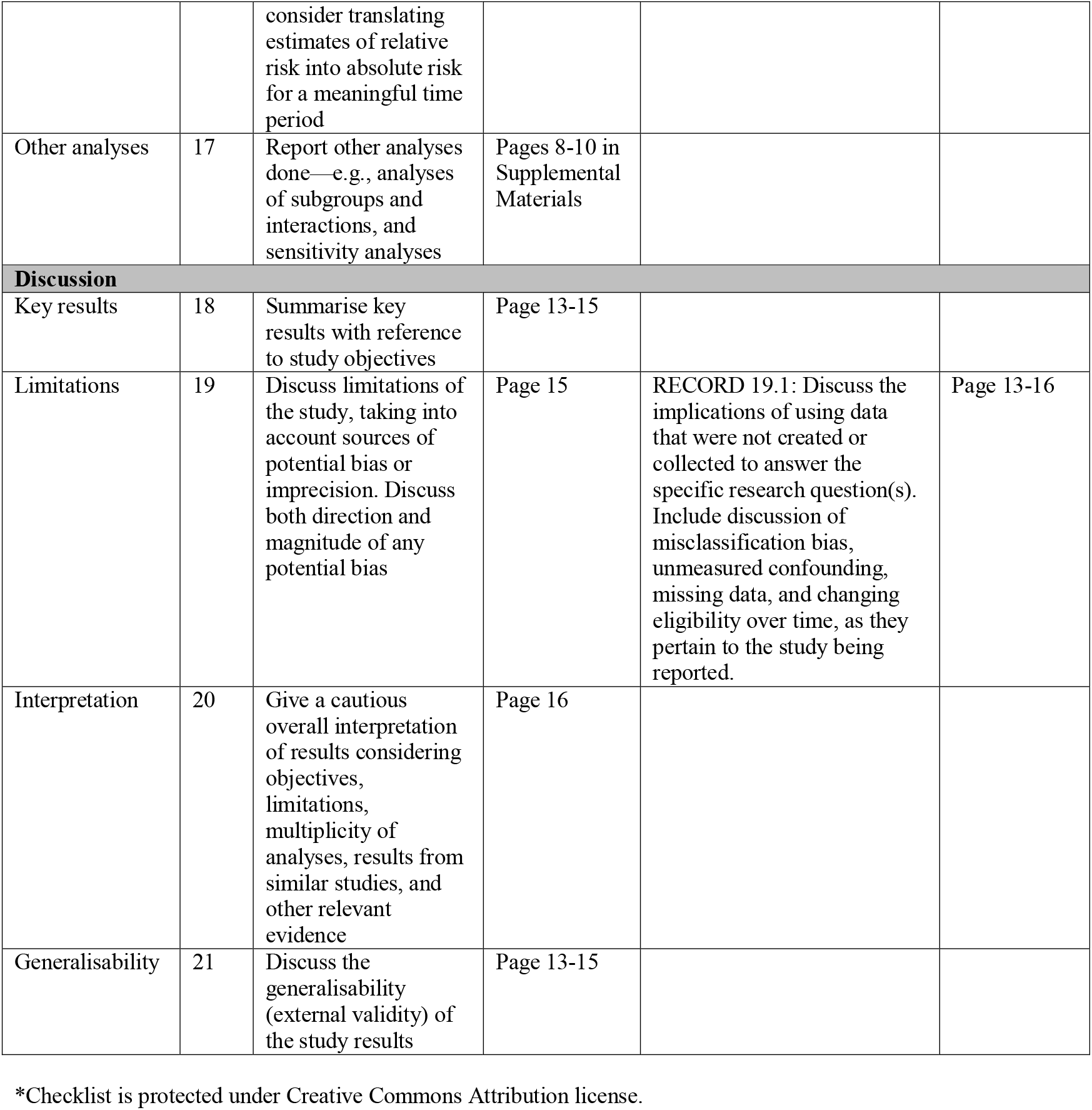
RECORD Statement Checklist

**eTable 2.**
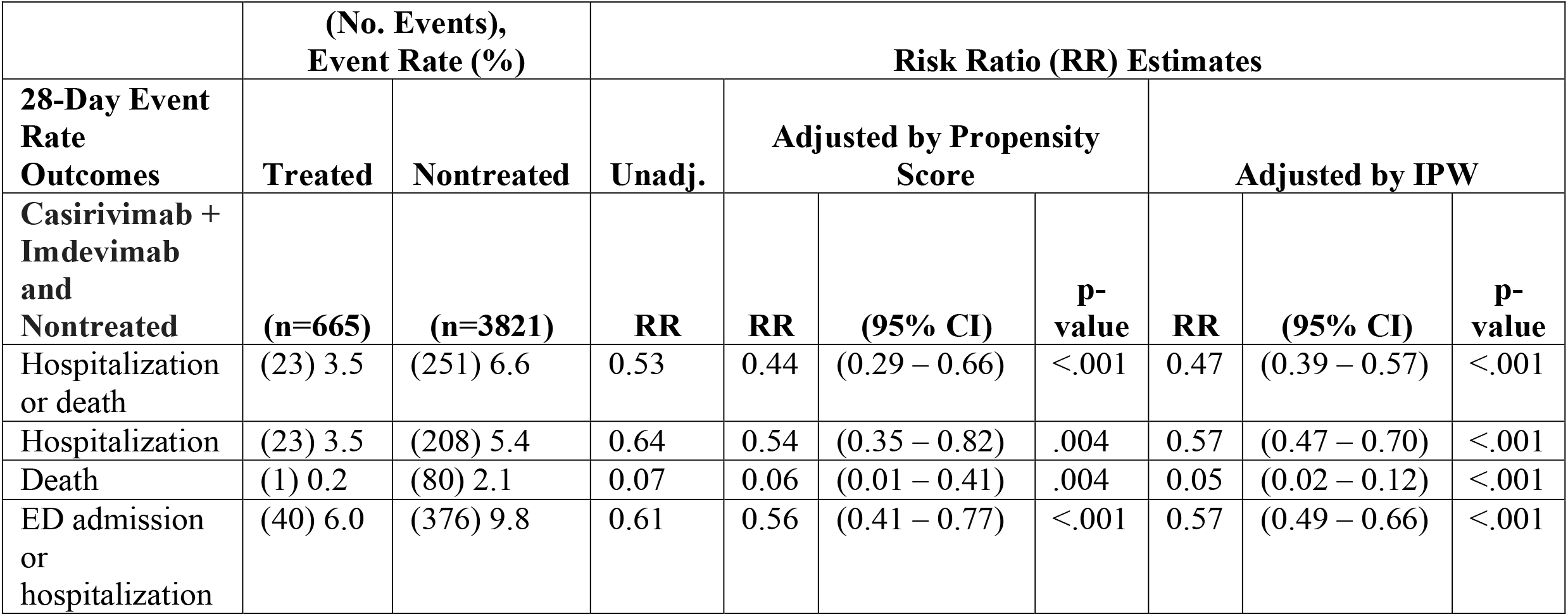
Primary and secondary outcomes in an unmatched cohort of patients receiving subcutaneous monoclonal antibody treatment and an at-risk population of patients not receiving monoclonal antibody treatment.

**eTable 3.** 28-Day Hospitalization Outcomes by Route of mAb Administration

| **Outcome** | **Subcutaneous**<br>**(No.), %** | **Intravenous**<br>**(No.), %** | **Absolute Risk**<br>**Difference (%)** |  |  |
| --- | --- | --- | --- | --- | --- |
|  |  |  | **Unadj** | **Adj.** | **95% CI** |
| **All Infused Patients\*** | **(n=969)** | **(n=1216)** |  |  |  |
| Hospitalization | (27), 2.8% | (20), 1.6% | 1.1% | 1.5% | -0.3% to 3.4% |
| ICU admission | (3), 0.3% | (3), 0.2% | 0.1% | 0.7% | -3.5% to 5.0% |
| Ventilation | (3), 0.3% | (2), 0.2% | 0.1% | 0.2% | -5.8% to 5.5% |
|  | **(n=27)** | **(n=20)** | **Wilcoxon p-value** |  |  |
| LOS of hospitalizations,<br>med, (IQR) | 4 (2,8) | 3 (4, 6.5) | 0.70 |  |  |
| **Same Site Infused**<br>**Patients\*\*** | **(n=721)** | **(n=441)** | **Unadj.** | **Adj.** | **95% CI** |
| Hospitalization | (17), 2.4% | (4), 0.9% | 1.5% | 1.5% | -0.4% to 3.7% |
| ICU admission | (3), 0.4% | (1), 0.2% | 0.2% | 0.2% | -5.9% to 6.3% |
| Ventilation | (2), 0.3% | (1), 0.2% | 0.1% | 0.0% | -8.7% to 8.6% |
|  | **(n=17)** | **(n=4)** | **Wilcoxon p-value** |  |  |
| LOS of hospitalizations,<br>med, (IQR) | 6 (3,10) | 4.5 (2, 6) | 0.42 |  |  |

\*Patients treated at all health system facilities; SQ patients were treated from July 20 – September 20, 2021; IV patients were treated from July 15- September 29, 2021. \*\*Patients treated at the same UPMC facilities; SQ patients were treated from September 9-29, 2021; IV patients were treated from July 15- September 29, 2021. Adj: Model adjusted for age, gender, and vaccination status.

**eFigure 1.**
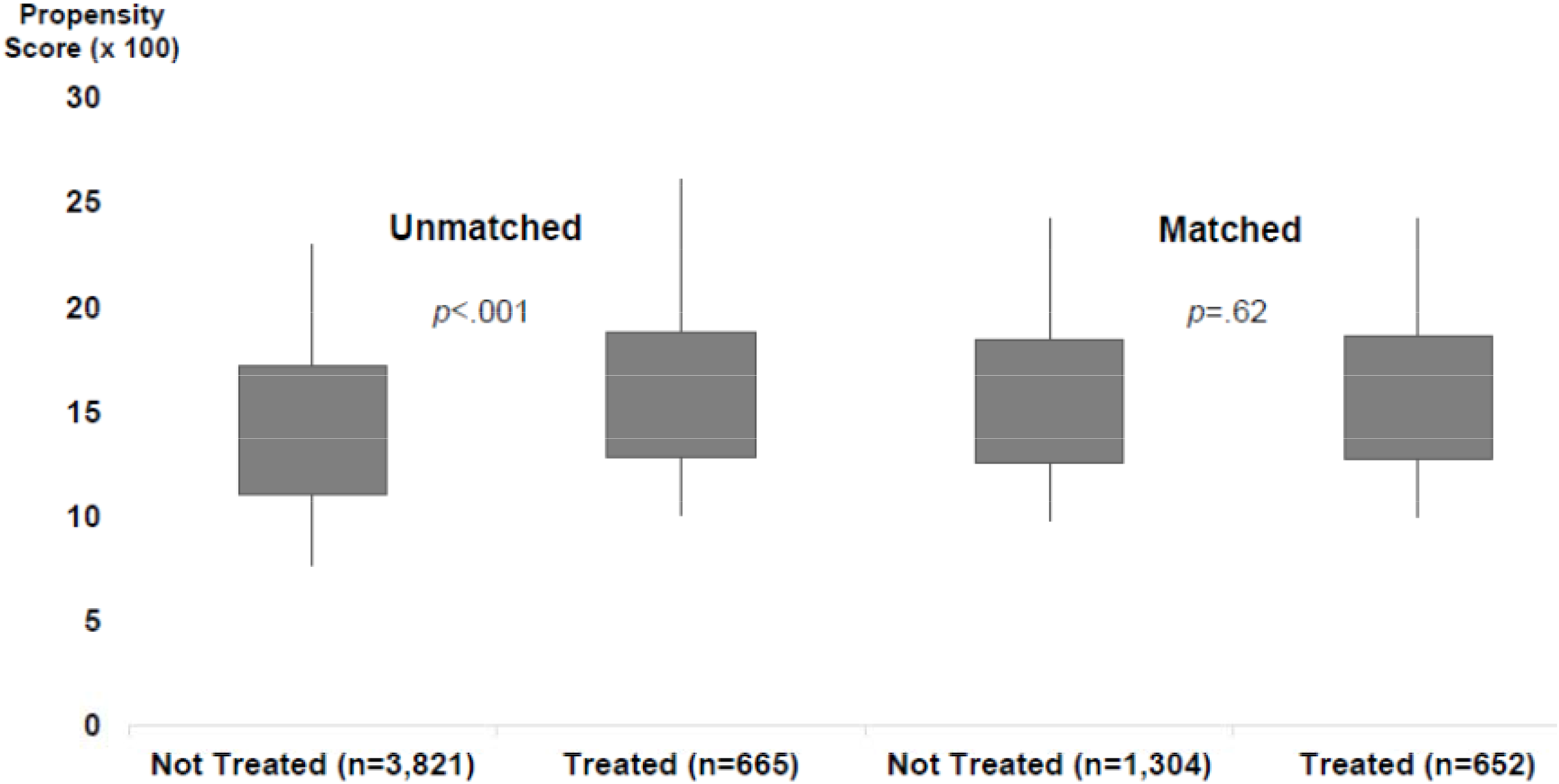
Distribution of propensity scores (x 100) before and after matching of treated and non-treated patients. The shaded rectangles depict the interquartile range; the lower and upper ends of the vertical lines depict the 5^th^ and 95^th^ percentiles. P-values are from Wilcoxon tests.

**eFigure 2.**
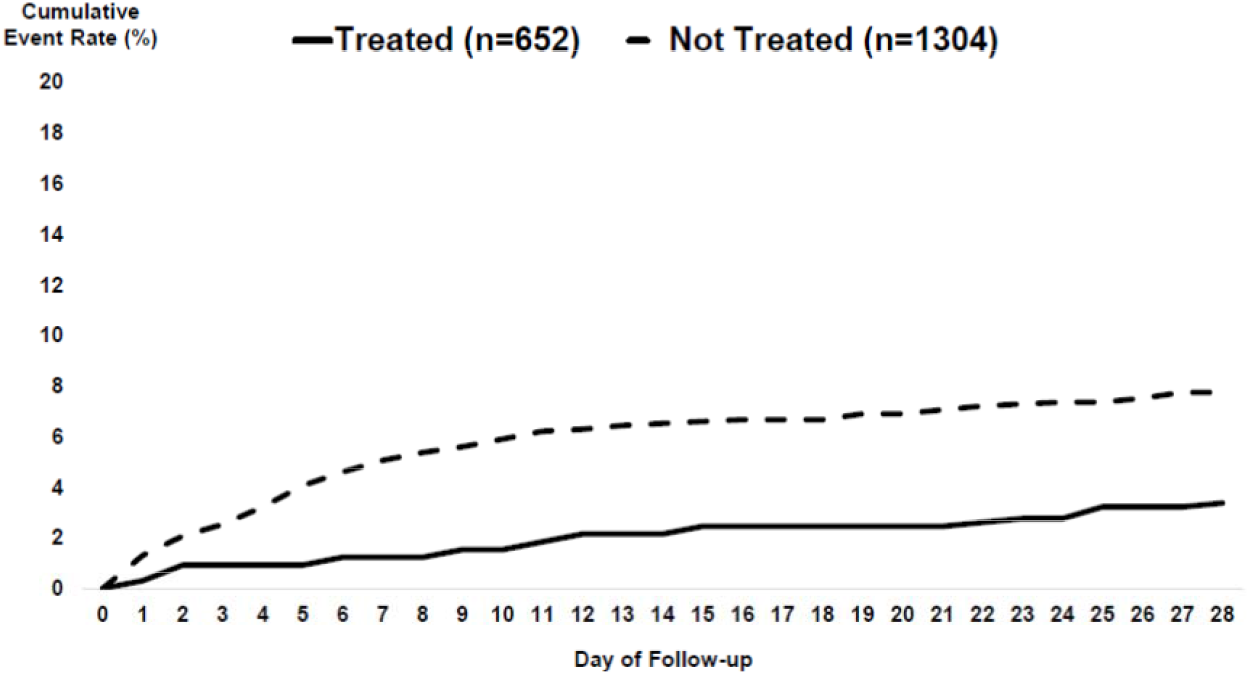
Plot of cumulative event rates of hospitalization/death by day of follow-up (x-axis) for matched treated (solid line) and non-treated (dashed line) patients.

## Notes

**Conflict of Interest:** None of the authors received any payments or influence from a third-party source for the work presented.

### Competing Interest Statement

The authors have declared no competing interest.

### Clinical Trial

NCT04790786

### Author Declarations

UPMC Quality Improvement Review Committee (Project ID 3629) and the University of Pittsburgh Institutional Review Board (STUDY21100151)gave ethical approval for this work.

